# Confirmatory insights into *ELMOD3*-associated autosomal dominant non-syndromic hearing loss

**DOI:** 10.1101/2025.02.11.25321773

**Authors:** Yejin Yun, Minjae Park, Sohyang Jeong, Sung Ho Jung, Sang-Yeon Lee

## Abstract

Sensorineural hearing loss (SNHL) is one of the most common sensory disorders, predominantly driven by monogenic causes with a higher Mendelian contribution. Although *ELMOD3* variants have been implicated in both autosomal dominant (DFNA81) and autosomal recessive (DFNB88) hereditary deafness, only a single DFNA81 family has been reported, leaving the pathogenic role of the dominant allele largely unexplored. Through targeted panel sequencing, we herein identified a novel heterozygous *ELMOD3* variant (c.640G>A; p.Gly214Ser) that co-segregates with autosomal dominant hearing loss in a Korean family. Molecular modeling and structure analysis indicates that replacing glycine at residue 214 with serine introduces spatial clashes with adjacent Ala160 and Cys162, thereby disrupting intermolecular interactions and compromising protein stability. Consistent with this, stability assays revealed a rapid degradation rate for the mutant protein.

Furthermore, the ability to localize with F-actin in mutant protein was disrupted compared to the wild-type protein. Based on functional assays, the p.Gly214Ser variant demonstrated the functional pathogenicity and was classified as likely pathogenic according to the ACMG guideline for hearing loss. Collectively, these findings provide confirmatory insights into *ELMOD3*-associated DFNA81, potentially through a dominant-negative mechanism.

## Introduction

Sensorineural hearing loss (SNHL) represents a prevalent sensory disorder with a higher Mendelian genetic contribution^1^. Recent advancements in the genetic understanding of SNHL have led to the identification of over 150 causative genes within human genetics^2,3^. Moreover, studies on mouse genetics have shed light on the physiological basis and molecular functions of SNHL in humans^4–7^. This genetic information has paved the way for the prediction of the natural progression of hearing loss via genotype-phenotype correlation, and is propelling the future development of inner-ear gene therapy to come. However, the clinical implications of genetic information regarding newly discovered or less-studied genes remain poorly understood. Consequently, reporting on such genes, including *ELMOD3*, which was initially associated with hearing loss in 2013 but has been reported in the literature in only two families so far, carries considerable importance^8,9^.

ELMOD3, a member of the ELMO family, is an ARL2 GTPase-activating protein^9^, and is known to play roles in cell adhesion, intracellular signaling, and the organization of actin cytoskeleton^9,10^. Defective Elmod3 have been associated with dysmorphologies in stereocilia *in vivo*, suggesting a potential role in the dynamics of the actin cytoskeleton within cochlear hair cells, which could be linked to hearing impairment^11^. The first variant (c.794T>C:p.Leu265Ser) was identified in a Pakistani family and caused non-syndromic hearing loss as an autosomal recessive trait (DFNB88)^9^. A second variant (c.512A>G:p.His171Arg) was identified in a Chinese family, with hearing loss in this case inherited as a dominant trait (DFNA81)^8^. The identification of *ELMOD3*-related hearing loss in humans and mice underscores the crucial role of *ELMOD3* in auditory function. However, the evidence linking *ELMOD3* variants to hearing loss in human genetics remains sparse, necessitating further studies to expand the genotype and associated phenotypes, as well as to unravel the molecular mechanisms of mutant proteins attributable to *ELMOD3* variants.

In this study, we identified a novel heterozygous *ELMOD3* variant (c.640G>A:p.Gly214Ser) that co-segregates with autosomal dominant hearing loss in a Korean family. To characterize this disease-causing variant, we conducted structural analysis and functional assays to investigate its pathogenic impact on *ELMOD3* function. Our findings contribute to the understanding of the causative role of *ELMOD3* in autosomal dominant hearing loss, providing further insight into the molecular basis of *ELMOD3*-associated DFNA81.

## Methods

### Participants

This study was a retrospective review using the in-house databases of Hereditary Hearing Loss Clinic within the Otorhinolaryngology division of the Center for Rare Diseases, Seoul National University Hospital, Korea. All study procedures were approved by the Institutional Review Boards of Seoul National University Hospital (IRB-H-0905-041-281). All participants/legal guardians were provided written informed consent to participation. In this study, one family segregating a heterozygous disease-causing variant of *ELMOD3* was included. The demographic data and clinical manifestations were retrieved from the electronic medical records.

### Targeted panel sequencing

DNA extraction from peripheral blood samples was carried out using the Chemagic 360 instrument (Perkin Elmer, Baesweiler, Germany). We used a SureSelect DNA targeted panel sequencing to capture the target regions, which incorporated 246 genes implicated in deafness (**Supplementary table 1**). The library was prepared and sequenced in a paired-end manner following the manufacturer’s guidelines, using an Illumina NovaSeq 6000 sequencing system (Illumina, San Diego, CA, USA). We aligned the sequence reads to the human reference genome (GRCh37) and adopted the Genome Analysis Toolkit (GATK) best-practice pipeline to identify single nucleotide variations (SNVs) and indels^12^. Annotation of variants was carried out using the ANNOVAR software, including the RefSeq gene set and the Genome Aggregation Database (gnomAD)^13,14^. Rare non-silent variants, including nonsynonymous SNVs, coding indels, and splicing variants, were selected as potential candidates. Ethnic-specific variants were further filtered through the Korean Reference Genome Database (KRGDB) and the Korean Variant Archive (KOVA2), databases dedicated to cataloging genetic variations in the Korean population^15,16^.

The ClinVar and HGMD databases were also screened to verify if the candidate variants had been identified in other patients previously. Sanger sequencing was employed to validate candidate variants, and parental DNA samples were used to evaluate phasing analysis. Finally, the pathogenicity classification of the candidate variants was determined according to the ACMG guideline for hearing loss^17^.

### Structure modeling

Based on the AlphaFold Protein Structure Database^18^, a homology model of the human ELMOD3 protein was developed. This model construction leveraged the ELMOD3 structure (AF-Q96FG2-F1) as a reference template. The impact of the *ELMOD3* p.Gly214Ser variant was evaluated by the Dynamut server (http://biosig.unimelb.edu.au/dynamut/) and the PyMOL software. We investigated intramolecular interactions (e.g., hydrophobic interactions, hydrogen bonds, and salt bridges) of the *ELMOD3* p.Gly214Ser variant to predict regional protein stability. All the graphical illustrations were produced using the PyMOL software (ver. 2.5.2) (PyMOL Molecular Graphics System ver. 2.0, Schrödinger Inc., New York, NY, USA).

### Expression plasmids and culture

A human *ELMOD3* cDNA clone (RC200857) was purchased from Origene. The *ELMOD3* p.Gly214Ser mutation plasmid was generated utilizing the QuickChange mutagenesis method^19^. HEI-OC1 cells were cultured in Dulbecco’s modified Eagle medium (DMEM) (LM001-05, Welgene), and MDCK cells were cultured in Minimum Essential Medium (MEM) (LM007-01, Welgene). All culture media were supplemented with heat-inactivated 10% fetal bovine serum, 100 units/mL penicillin/streptomycin, and 2 mM L-glutamine. The MDCK cells were maintained in humidified air with 5% CO2 at 37°C. For western blotting, cells were transfected with 2 ug of total plasmid DNA in a 6-well plate using Lipofectamine 3000 Reagent (L3000015, Thermo), following the manufacturer’s protocols. For immunocytochemistry, MDCK cells were transfected with 0.5 ug of total plasmid DNA in a 24-well plate using Fugene HD Transfection Reagent (E5911, Promega), following the manufacturer’s protocols.

### Immunocytochemistry

Cultured cells on cover glasses were incubated for 20 minutes with 4% paraformaldehyde. After washing three times with PBS, fixed cells were permeabilized and blocked with PBS containing 0.5% Triton X-100 and 1% BSA for 20 minutes at room temperature. Then, the cells were incubated overnight with primary antibody at 4°C and washed three times with 0.1% Tween-20 in PBS. Afterward, cells were incubated for 30 minutes with secondary antibody conjugated with Alexa Fluor 488 at room temperature. After washing three times, cells were mounted on a glass slide with an antifading mounting medium containing 4’6’-diamidino-2-phenylindole (DAPI). Images were taken with a laser scanning confocal microscope (Leica STELLARIS 8, Upright).

### Western blot

Transfected HEK293T cells were lysed using RIPA lysis buffer containing protease inhibitor. After being incubated for 15 minutes at 4°C, whole-cell lysates were centrifuged at 13000 rpm for 15 minutes at 4°C. Then, the supernatant was acquired and mixed with 4X LDS sample buffer. Separation of proteins in whole-cell lysates was done using 12% sodium dodecyl sulfate-polyacrylamide gel electrophoresis (SDS-PAGE), and proteins were transferred to a polyvinylidene difluoride (PVDF) membrane (IPVH00010, Millipore). The membrane was incubated overnight with primary antibody at 4°C. After washing three times with TBS-T, the membrane was incubated for 1 hour with secondary antibody conjugated with horseradish peroxidase at room temperature. After washing three times, protein bands were detected with enhanced chemiluminescence.

### Cycloheximide chase assay

To identify the stability of ELMOD3 proteins, HEI-OC1 cells were transfected with 1 ug of total plasmid DNA in a 12-well plate using Lipofectamine 3000 Reagent (L3000015, Thermo), following the manufacturer’s guidelines. pCMV6-ELMOD3-Myc-Flag and pCMV-ELMOD3 p.Gly214Ser-Myc-Flag plasmids were transfected to 6 wells of the 12-well plate, respectively. After 24 hours, 80 μg/μl of cycloheximide was treated and cells were harvested after 0, 2, 4, 6 and 8 hours. Western blot of each sample was performed as described above, in the Western blot section. For quantification, ELMOD3 expression was compared with β-actin for each sample using densitometry. ImageJ software was utilized to analyze the obtained data and calculate the degradation rate.

## Results

### Clinical phenotype

In Family 1, a familial history of hearing impairment was identified, segregating as a dominant trait, with Sanger sequencing confirming heterozygous variants in the affected individuals (**Fig.1A**). The age of onset for hearing loss for the proband (Family 1-1) was the late 20s, respectively. The proband exhibited progressive deterioration in hearing, and the audiogram showed symmetric moderately-severe SNHL with a down-sloping configuration (**Fig.1B**). Neither temporal bone computed tomography (CT) nor internal acoustic canal magnetic resonance imaging (MRI) revealed any inner ear anomalies of the proband. A review of the medical history and physical examination of the proband did not show other symptoms or underlying disease. The auditory phenotypes for Family 1 members who were available for genotyping are summarized (**Supplementary Table 2**).

**Figure 1.**
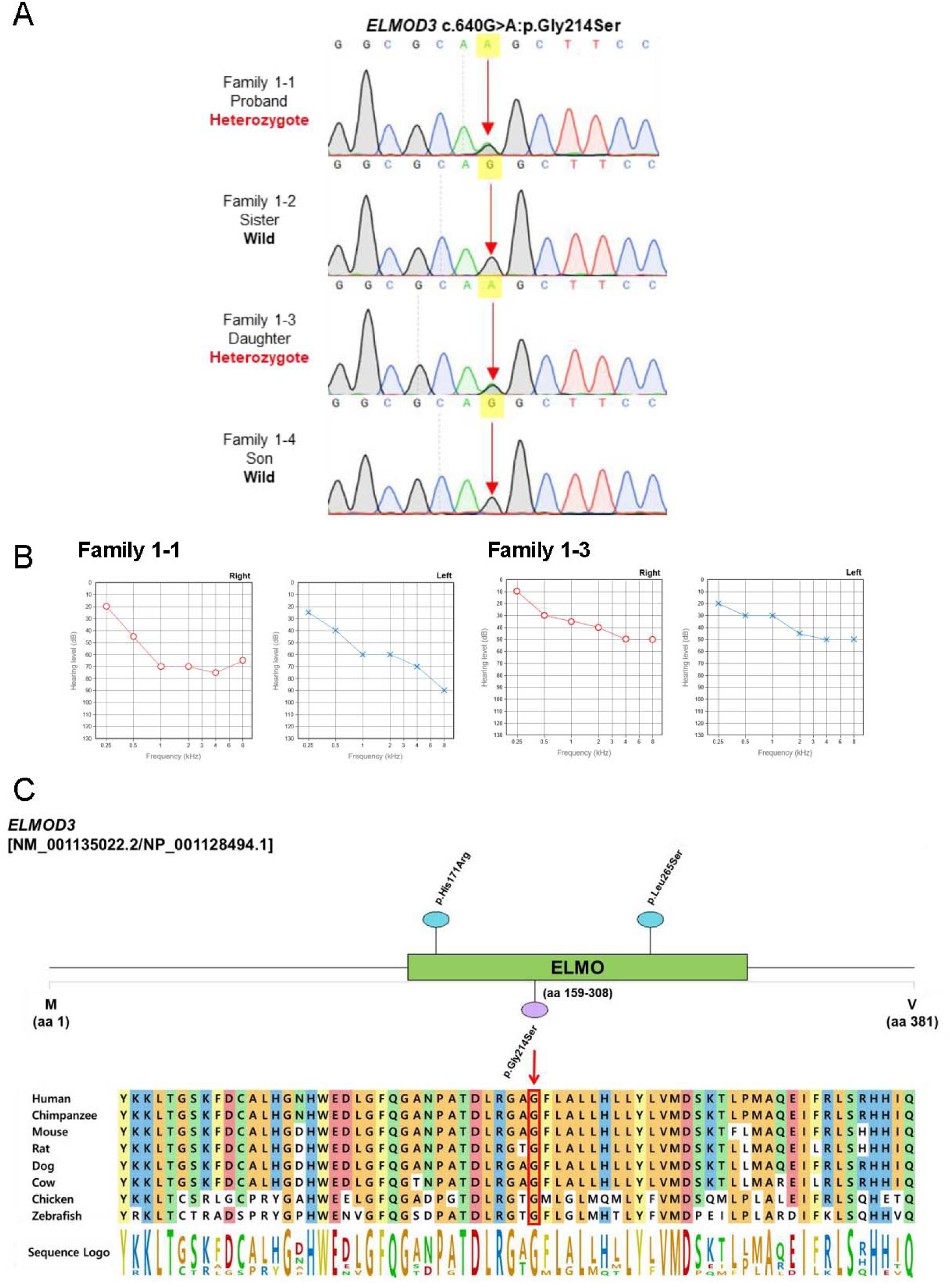
Clinical phenotypes and genotype for *ELMOD3* novel variant-associated with autosomal dominant hearing loss. (A) Results of segregation analysis by Sanger sequencing. (B) Pure tone audiograms of the affected individuals depicted in the pedigree. (C) Protein domain and conservation map of ELMOD3. Previously reported variants and the novel variant are located at the ELMO domain. The Gly214 residue, denoted with an arrow, is conserved between species.

### Genotype

Through targeted panel sequencing, we identified a novel heterozygous variant (c.640G>A:p.Gly214Ser) in *ELMOD3* underlying autosomal dominant non-syndromic hearing loss. In the application of a next-generation sequencing (NGS)-based copy number variation (CNV) detection algorithm, specifically the CCR-CNV analysis^20^, we observed no evidence of deletions or duplications in the exomes of the 246 deafness genes.

The novel variant p.Gly214Ser of *ELMOD3* is noted as the third reported mutant allele associated with SNHL and represents the second mutant allele of an autosomal dominant trait. The variant is absent across various genomic databases, including the KRGDB/KOVA and the gnomAD database. In silico analyses revealed that this missense variant had a deleterious impact, scoring 29.1 and 0.843 on the Combined Annotation Dependent Depletion (CADD) and Rare Exome Variant Ensemble Learner (REVEL) algorithms, respectively. The amino acid residue at the Gly214 position is highly conserved across orthologous genes of various species and has a higher GERP score of 5.76, which underlines its possible functional significance. Sanger sequencing, employed for phasing analysis, demonstrated that the identified variant co-segregated with the hearing loss phenotype in individuals. According to the ACMG guideline for hearing loss, the p.Gly214Ser variant was initially classified as VUS since it meets the criteria for PM2, PP1_Moderate, and PP3. Based on functional assays, the p.Gly214Ser variant demonstrated the functional pathogenicity (PS3_supporting) and was consequently classified as likely pathogenic.

### Molecular modeling and structure analysis

The structure of the ELMOD3 model generated by AlphaFold (AF-Q96FG2-F1) is depicted in **Fig.2A**. We used this AlphaFold-generated model structure of ELMOD3 to investigate the impacts of the Gly214Ser missense variant on the ELMOD3 protein structure, in comparison to the wild-type protein. The Gly214 amino acid residue is located in the helix of the ELMO domain. Upon the substitution of glycine at position 214 with serine, spatial clashes with neighboring Ala160 and Cys162 residues occur, which affect intermolecular interactions with backbone molecules such as hydrogen bonding (**Fig.2B**). There are three possible rotamer states, specifically R1, R2, and R3, resulting from the substitution of Ser214. The collisions with neighboring amino acid residues occur in any of these rotamer states.

**Figure 2.**
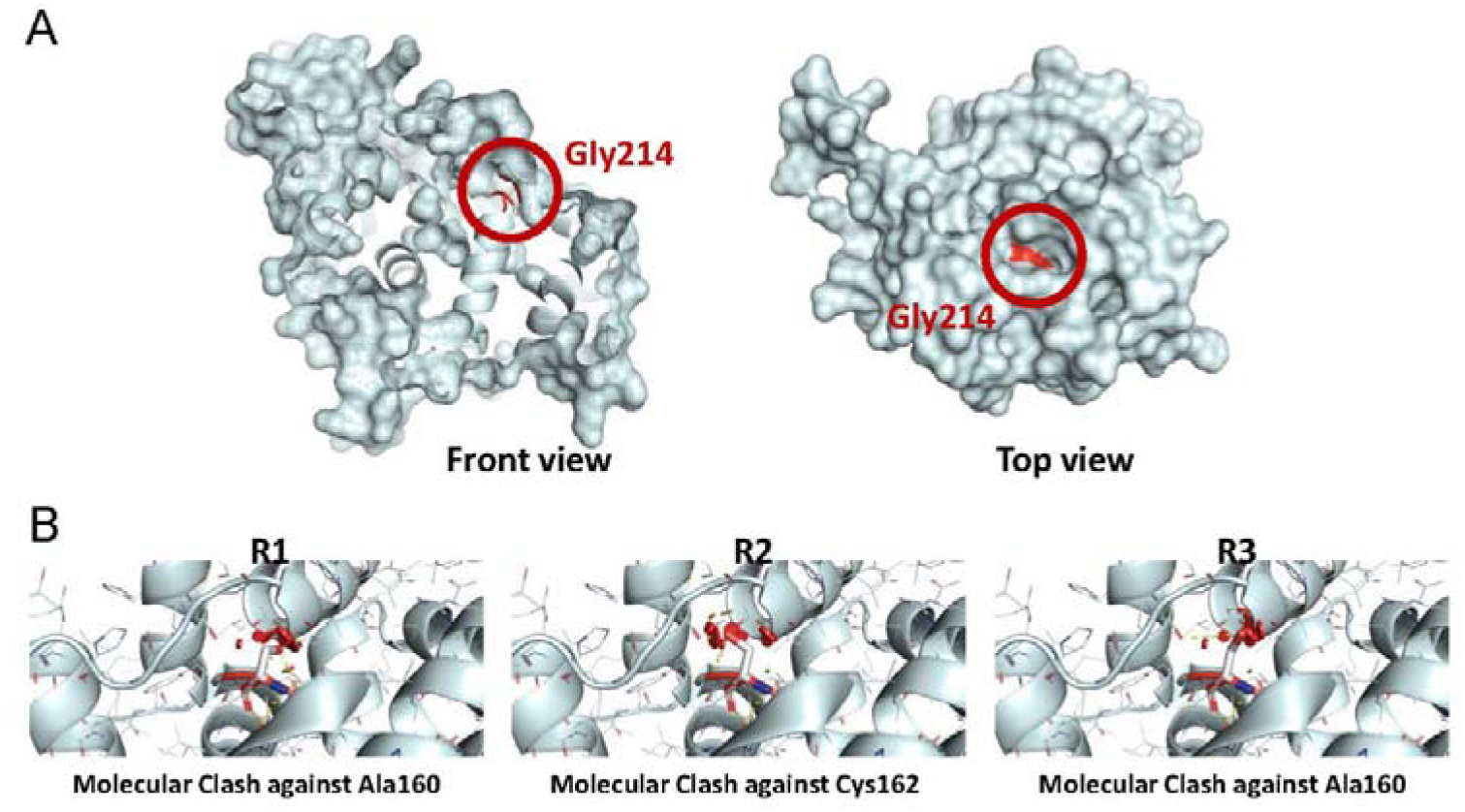
AlphaFold-based molecular modeling of ELMOD3 and prediction of the variant’s impact on molecular structure. (A) Molecular model of ELMOD3 generated via AlphaFold. Gly214 is located at the helix of the ELMO domain. (B) Molecular clashes occurred by the p.Gly214Ser variant, affecting intermolecular interactions. The variant leads to spatial clashes with adjacent amino acid residues in every possible rotamers, predicted to affect stability of the protein.

Collectively, the identified *ELMOD3* variant compromises protein stability.

### Subcellular localization

To confirm whether the ELMOD3 mutant protein interrupts with subcellular localization, we conducted an immunofluorescence study. MDCK cells were transfected with pCMV6-ELMOD3-Myc-Flag and pCMV-ELMOD3 p.Gly214Ser-Myc-Flag plasmids. To identify subcellular distribution of proteins, antibodies were used to detect ELMOD3 and F-actin. Wild type ELMOD3 expressed in the cytoplasm and plasma membrane (**Fig. 3A**). However, the mutant protein was detected abundantly in the nucleus, with sparse expression in the cytoplasm (**Fig. 3B**). Co-localization with F-actin, which was expressed in the cytoplasm and plasma membrane, was significantly decreased in the mutant protein. Hence, the mutant’s ability to localize with F-actin was disrupted compared to the wild-type protein.

**Figure 3.**
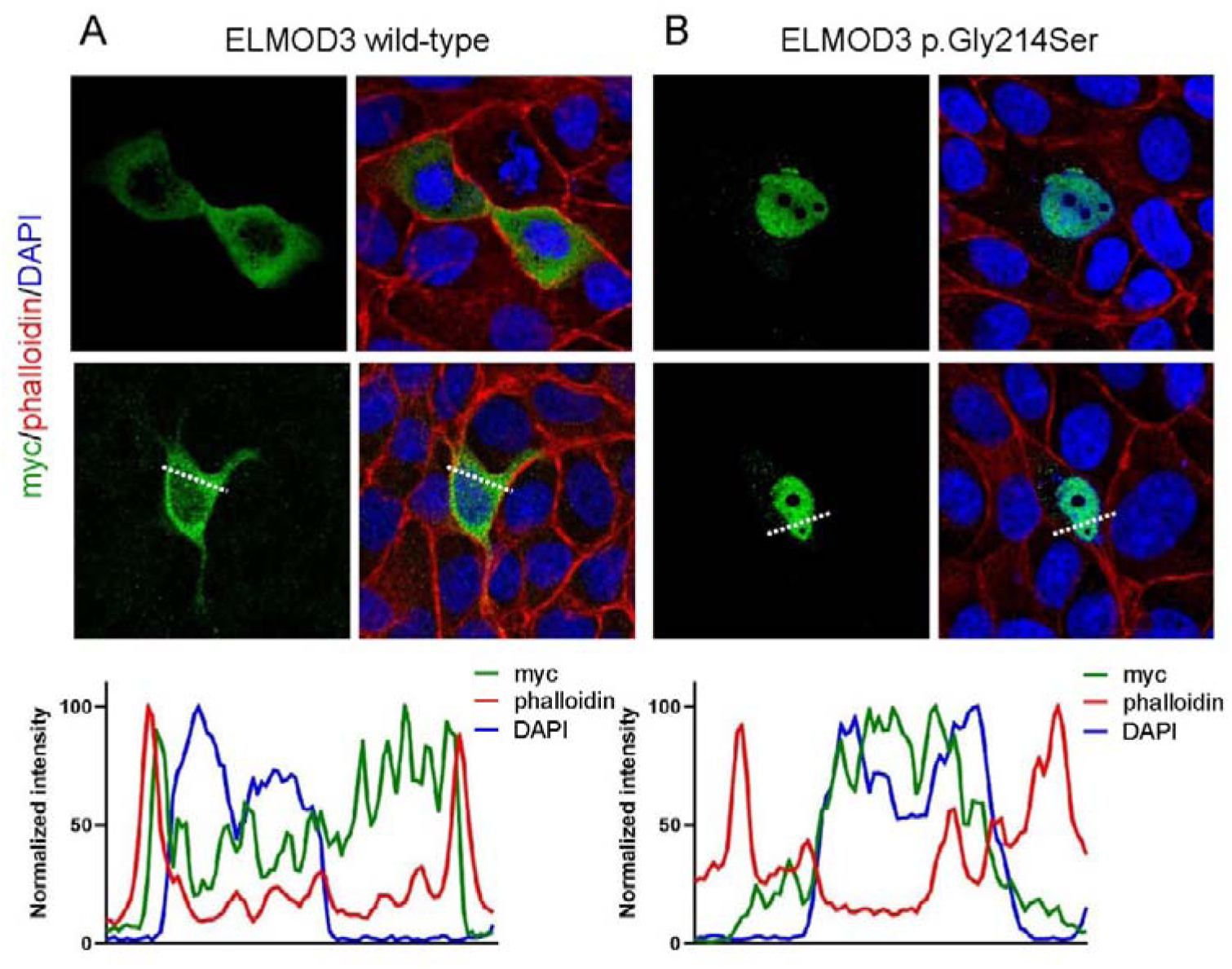
Effects of *ELMOD3* p.Gly214Ser variant to localize with F-actin. MDCK cells were transfected with pCMV6-ELMOD3-Myc-Flag and pCMV-ELMOD3 p.Gly214Ser-Myc-Flag. ELMOD3 was detected using anti-myc antibody (green) and F-actin by phalloidin (red). (A) In the wild-type, ELMOD3 was detected in the cytoplasm and plasma membrane, co-localizing with F-actin. (B) In the mutant protein, co-localization of ELMOD3 with F-actin was disrupted, with abundant expression in the nucleus.

### Protein expression

Western blot assay using HEI-OC1 cells was performed to identify the impact of p.Gly214Ser mutation on ELMOD3 protein size and expression level. HEI-OC1 cells were transfected with pCMV6-ELMOD3-Myc-Flag and pCMV-ELMOD3 p.Gly214Ser-Myc-Flag plasmids. Wild-type and mutant ELMOD3 were both detected with same size, and no significant difference was shown considering expression levels (**Fig.4A**).

**Figure 4.**
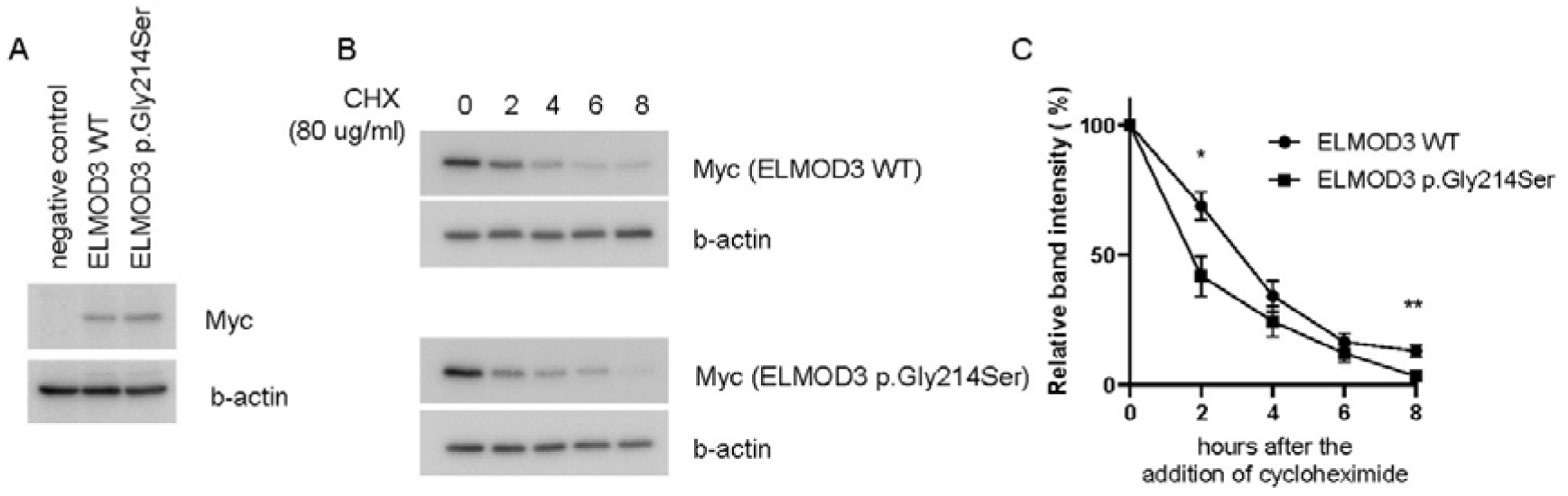
Effects of *ELMOD3* p.Gly214Ser variant on protein stability. (A) Protein expression levels of ELMOD3 WT and p.Gly214Ser mutant. (B) Cycloheximide was added to HEI-OC1 cells, which was transfected with pCMV6-ELMOD3-Myc-Flag and pCMV-ELMOD3 p.Gly214Ser-Myc-Flag. After 0, 2, 4, and 8 hours, cells were lysed, and Western blot was performed for each sample. Expression levels of ELMOD3 were compared using β-actin as an internal reference. (C) Relative abundance of wild type and mutant ELMOD3 by hours after cycloheximide treatment. At 2 and 8 hours, relative expression of mutant was decreased compared to wild type.

### Protein stability

To further verify the prediction of molecular modeling prediction, we conducted a stability assay using cycloheximide. HEI-OC1 cells transfected with *ELMOD3* were harvested 0, 2, 4, 6, and 8 hours after cycloheximide treatment and undergone Western blot assay (**Fig.4B**). Using β-actin as an internal control, relative protein abundance was analyzed. Compared to the wild type, the relative expression level of ELMOD3 was significantly decreased in the mutant at 2 and 8 hours, implicating an increased degradation rate of mutant protein (*p* = 0.028 and *p* = 0.0072, respectively) (**Fig.4C**). In short, the stability of ELMOD3 protein was compromised by the p.Gly214Ser variant, leading to its rapid degradation.

### Literature review and genotype-phenotype correlation

To date, including our study, three variant alleles of *ELMOD3* have been reported to be linked to hearing loss in human (**Table 1**). All three variants are coding variants located in the engulfment and cell motility (ELMO) domain (**Fig.1C**). The first variant, c.794T>C, was identified in consanguineous Pakistani family. The mutation was inherited in an autosomal recessive pattern, with the phenotype of prelingual, severe-to-profound mixed hearing loss. The second variant, c.512G>A, was identified in a Chinese family, which is inherited in an autosomal dominant pattern. Deaf individuals in the family expressed post-lingual, slowly progressive, severe-to-profound SNHL with the average onset of 28.3 years. In this study, we identified the variant of c.640G>A from a South Korean family as the third variant linked to hearing loss, which is inherited in an autosomal dominant pattern. Reported cases of *ELMOD3*-related DFNA81 expressed hearing loss phenotypes of symmetry, post-lingual onset SNHL (**Table 2**). No symptoms other than hearing loss were present in all reported cases.

**Table 1.** *ELMOD3* variants associated with hearing loss and its pathogenicity.

| Genomic Position<br>(GRCh37/hg19) | HGVS |  | Zygosity | Location<br>(Exon/Domain) | Insilico Predictions |  |  | Allele Frequency |  | ACMG 2018 | Reference |
| --- | --- | --- | --- | --- | --- | --- | --- | --- | --- | --- | --- |
|  | Nucleotide<br>change | Amino Acid<br>change |  |  | CADD<br>Phred | REVEL | Splice AI /<br>MAXENT<br>Score | KRGDB/<br>KOVA | gnomAD | Classification |  |
| chr2:g.85616929<br>T>C | c.794T>C | p.Leu265Ser | Homozygote | exon 12 / ELMO | 29.10 | 0.53 | AG(0.02)<br>DG(0.01)<br>/ NA | absent | 0.000003978<br>(1/251362) | Pathogenic | Jaworek et al.<br>(2013) |
| chr2:g.85598590<br>A>G | c.512A>G | p.His171Arg | Heterozygote | exon 10 / ELMO | 26 | 0.693 | AG(0.08)<br>DG(0.07)<br>/ 5'ss(5.65) | absent | 0.000007958<br>(2/251314) | Pathogenic | Li et al. (2018) |
| chr2:g.85604499<br>G>A | c.640G>A | p.Gly214Ser | Heterozygote | exon 11 / ELMO | 29.7 | 0.843 | (0.00) / NA | absent | 0.000004057<br>(1/246506) | Likely<br>Pathogenic | This study |

Refseq transcript accession number NM\_001135022.2; Refseq protein accession number NP\_001128494.1.
HGVS: Human Genome Variation Society (https://www.hgvs.org/); Sequence Variant Nomenclature (http://varnomen.hgvs.org/); CADD: Combined Annotation Dependent Depletion (https://cadd.gs.washington.edu/); REVEL: Rare Exome Variant Ensemble Learner (https://sites.google.com/site/revelgenomics/); KRGDB: Korean Reference Genome Database (http://coda.nih.go.kr/coda/KRGDB/index.jsp); gnomAD: The Genome Aggregation Database (https://gnomad.broadinstitute.org/); ACMG/AMP 2018 guideline (http://wintervar.wglab.org/). ELMO indicates Engulfment and cell motility.

**Table 2.** Clinical phenotypes of previously reported families with *ELMOD3*-associated hearing loss.

| Family ID | Age/Sex | Gene | Variants | Zygosity | Clinical phenotypes |  |  |  | References |
| --- | --- | --- | --- | --- | --- | --- | --- | --- | --- |
|  |  |  |  |  | onset | severity | asymmetry | progressive |  |
| Family 2 | 21-25/F | *ELMOD3* | c.794T>C | Homozygote | prelingual | profound | - | - | Jaworek, et al. (2013) |
|  | 16-20M | *ELMOD3* | c.794T>C | Homozygote | prelingual | profound | - | - | Jaworek, et al. (2013) |
| Family 3 | 31-35/M | *ELMOD3* | c.512A>G | Heterozygote | postlingual | moderate | - | progressive | Li, et al. (2018) |
|  | 36-40/F | *ELMOD3* | c.512A>G | Heterozygote | postlingual | severe | - | progressive | Li, et al. (2018) |
|  | 46-50/F | *ELMOD3* | c.512A>G | Heterozygote | postlingual | N/A | N/A | progressive | Li, et al. (2018) |
|  | 51-55/M | *ELMOD3* | c.512A>G | Heterozygote | postlingual | profound | - | progressive | Li, et al. (2018) |
|  | 56-60/M | *ELMOD3* | c.512A>G | Heterozygote | postlingual | profound | - | progressive | Li, et al. (2018) |

*ELMOD3*, transcript NM\_001135022.2; Abbreviations: N/A, not available

## Discussion

In this study, we discovered a novel *ELMOD3* variant leading to dominant form of hearing loss via targeted panel sequencing. Molecular modeling and structure analysis indicates that replacing glycine at residue 214 with serine introduces spatial clashes with adjacent Ala160 and Cys162, thereby disrupting intermolecular interactions and compromising protein stability. Consistent with this, stability assays revealed a rapid degradation rate for the mutant protein. Furthermore, the ability to localize with F-actin in mutant protein was disrupted compared to the wild-type protein. Based on functional assays, the p.Gly214Ser variant demonstrated the functional pathogenicity and was classified as likely pathogenic according to the ACMG guideline for hearing loss.

Proteins in the family of ELMODs are known to act as a GTPase activating protein (GAP) against Arf-family small GTP binding proteins. In specific, ELMOD3 exhibited GAP activity against ARL2, which is known to be associated with the regulation of actin cytoskeleton dynamics via intracellular signaling pathways. A recessive variant of *ELMOD3* was associated with reduction of ARL2 GTPase activity, *in vitro.*^9^ Moreover, in a recent study, it was proposed that lack of ELMOD3 was associated with improper transportation of proteins to cilia, resulting in reduction of ciliation^21^. In hair cells of developing rat cochlea, ELMOD3 expression was detected in stereocilia, kinocilia, and cuticular plate, implicating its role in auditory sensory cells. Also, *in vivo* study on *Elmod3* knock-out mice have shown that the lack of Elmod3, which leads to reduction of ARL2 expression, is associated with morphological dysgenesis of stereocilia within murine cochlear hair cells^11^. The failure of p.Gly214Ser variant to localize with actin cytoskeleton, and its altered degradation kinetics is expected to deteriorate actin cytoskeleton in hair cell stereocilia, leading to its dysmorphologies. The change of stereocilia morphology within cochlear hair cells alters their function, being responsible for hearing loss in affected individuals.

Authors of a previous study on dominant form of hearing loss caused by an *ELMOD3* variant have insisted that haplo-insufficiency is sufficient to explain the phenotype caused in heterozygotes^8^. However, *in vivo* study using *Elmod3* knock-out mice, heterozygous murine models were not associated with any phenotypes^11^. In addition, the probability of loss-of-function intolerance (pLI) score of *ELMOD3* calculated from gnomAD were 0.00, suggesting that its susceptibility to haplo-insufficiency is extremely low^14^. Based on this, we suggest that dominant-negative mechanism may underlie *ELMOD3*-associated DFNA81.

In conclusion, we identified a novel *ELMOD3* variant (c.640G>A; p.Gly214Ser) linked to autosomal dominant form of hearing loss (DFNA81). Structural modeling and functional assay revealed that a novel heterozygous *ELMOD3* variant compromise protein stability and disrupted localization with F-actin. Collectively, these findings provide additional evidence of *ELMOD3*-associated DFNA81, potentially through a dominant-negative mechanism.

## Data Availability

The data generated in this study, is available from the corresponding author upon request.

---

**Supplementary table 1.**
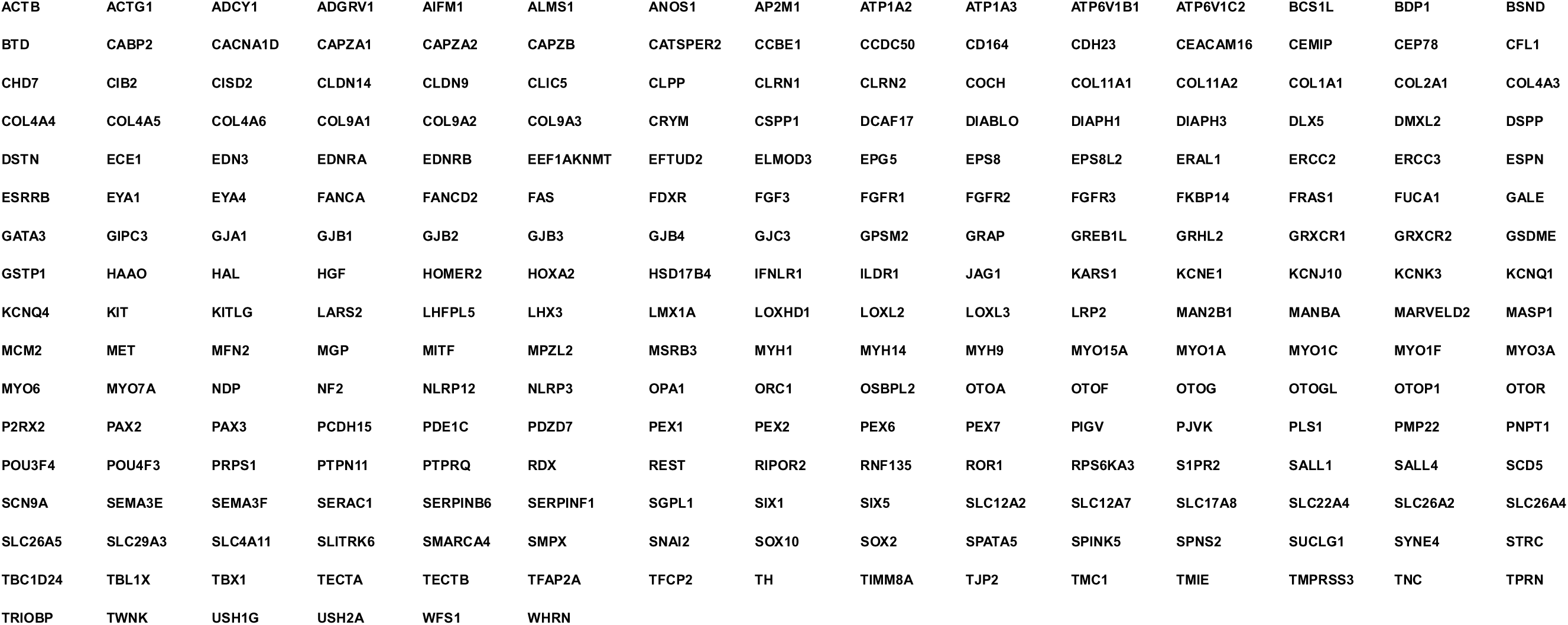
Gene List for targeted panel sequencing.

**Supplementary table 2.** Clinical phenotypes of the affected and nonaffected individuals in the Family 1.

| Family ID | Age/Sex | Gene | Variants | Zygosity | Clinical phenotypes |  |  |  |
| --- | --- | --- | --- | --- | --- | --- | --- | --- |
|  |  |  |  |  | onset | severity | asymmetry | progressive |
| Family 1-1 | 71-75/F | *ELMOD3* | c.640G>A | Heterozygote | postlingual | moderately severe | - | progressive |
| Family 1-3 | 46-50//F | *ELMOD3* | c.640G>A | Heterozygote | postlingual | mild-to-moderate | - | N/A |
| Family 1-2 | 66-70/F | *ELMOD3* | Wild type | - | - | - | - | - |
| Family 1-4 | 51-55/M | *ELMOD3* | Wild type | - | - | - | - | - |

*ELMOD3*, transcript NM\_001135022.2; Abbreviations: N/A, not available

